# Epidemiological and clinical features of 291 cases with coronavirus disease 2019 in areas adjacent to Hubei, China: a double-center observational study

**DOI:** 10.1101/2020.03.03.20030353

**Authors:** Xu Chen, Fang Zheng, Yanhua Qing, Shuizi Ding, Danhui Yang, Cheng Lei, Zhilan Yin, Xianglin Zhou, Dixuan Jiang, Qi Zuo, Jun He, Jianlei Lv, Ping Chen, Yan Chen, Hong Peng, Honghui Li, Yuanlin Xie, Jiyang Liu, Zhiguo Zhou, Hong Luo

## Abstract

**Background:** The clinical outcomes of COVID-19 patients in Hubei and other areas are different. We aim to investigate the epidemiological and clinical characteristics of patient with COVID-19 in Hunan which is adjacent to Hubei.

**Methods:** In this double-center, observational study, we recruited all consecutive patients with laboratory confirmed COVID-19 from January 23 to February 14, 2020 in two designated hospitals in Hunan province, China. Epidemiological and clinical data from patients’ electronic medical records were collected and compared between mild, moderate and severe/critical group in detail. Clinical outcomes were followed up to February 20, 2020.

**Findings:** 291 patients with COVID-19 were categorized into mild group (10.0%), moderate group (72.8%) and severe/critical group (17.2%). The median age of all patients was 46 years (49.8% were male). 86.6% patients had an indirect exposure history. The proportion of patients that had been to Wuhan in severe/critical group (48.0% vs 17.2%, p=0.006) and moderate group (43.4% vs 17.2%, p=0.007) were higher than mild group. Fever (68.7%), cough (60.5%), and fatigue (31.6%) were common symptoms especially for severe and critical patients. Typical lung imaging finding were bilateral and unilateral ground glass opacity or consolidation. Leukopenia, lymphopenia and eosinopenia occurred in 36.1%, 22.7% and 50.2% patients respectively. Increased fibrinogen was detected in 45 of 58 (77.6%) patients with available results. 29 of 44 (65.9%) or 22 of 40 (55.0%) patients were positive in Mycoplasma pneumonia or Chlamydia pneumonia antibody test respectively. Compared with mild or moderate group, severe/critical group had a relative higher level of neutrophil, Neutrophil-to-Lymphocyte Ratio, h-CRP, ESR, CK, CK–MB, LDH, D-dimer, and a lower level of lymphocyte, eosinophils, platelet, HDL and sodium (all p<0.01). Most patients received antiviral therapy and Chinese Medicine therapy. As of February 20, 2020, 159 (54.6%) patients were discharged and 2 (0.7%) patients died during hospitalization. The median length of hospital stay in discharged patients was 12 days (IQR: 10-15).

**Interpretation:** The epidemiological and clinical characteristics of COVID-19 patients in Hunan is different from patients in Wuhan. The proportion of patients that had been to Wuhan in severe/critical group and moderate group were higher than mild group. Laboratory and imaging examination can assist in the diagnosis and classification of COVID-19 patients.

## 1. Introduction

In December 2019, Coronavirus disease 2019 (COVID-19) broke out in Wuhan, China, and quickly spread to other Chinese provinces and 38 countries around the world up to February 26, 2020. Owing to its involvement in multiple areas, WHO declared a Public Health Emergency of International Concern on January 30, 2020. Up to February 26, 2020, there were 78,630 laboratory-confirmed and clinical-confirmed cases in China and over 3,000 cases outside China while about 2/3 of cases in China was located in Wuhan.^1^

Zhang et al reported the largest epidemiological investigation of 72314 cases and showed the crude case fatality rate in Hubei province (2.9%) was 7.3 times higher than other provinces (0.4%) in China,^2^ which indicated that there are differences in clinical outcome between the patients in Hubei and other provinces. The features and outcomes of patients with COVID-19 in Wuhan have been described in detail in several studies. Currently, there has been limited studies about patients with COVID-19 in Zhejiang and Beijing, which is far from Hubei.^3,4^ However, the clinical characteristics and progression of disease outside Hubei, especially in areas near Hubei which have a relative higher risk of importing patients than remote areas, were still unknown.

Hunan is adjacent to Hubei province, and the well-developed transportation system between Hunan and Hubei provided a high possibility for disease transmission in the early stage of COVID-19 outbreak when measures like city lockdown and traffic restriction were not taken. Therefore, studies in areas near Hubei can provide more information about the clinical characteristics of COVILD-19 and experience of diagnosis and treatment which can be referenced by areas outside Wuhan and countries worldwide under the current epidemic of COVID-19. In this study, we aimed to investigate the epidemic history and clinical characteristics of patients with COVID-19 in Hunan, China.

## 2. Methods

### 2.1 Study design and subjects

From January 23 to February 14, 2020, all consecutive patients with confirmed COVID-19, admitted to the first Hospital of Changsha and Loudi Central Hospital in Hunan, China were recruited. The first Hospital of Changsha and Loudi Central Hospital are two of the major tertiary hospitals and are responsible for the treatments for patients with COVID-19 assigned by the Chinese government. Diagnostic criteria and clinical classification of all confirmed COVID-19 cases were based on guidelines of National Health Commission.^5^ Data collection and analysis of cases and close contacts were determined by the National Health Commission of the People’s Republic of China (PRC) to be part of a continuing public health outbreak investigation and were thus considered exempt from institutional review board approval. Oral consent was obtained from all patients.

### 2.2 Data collection

Epidemiological, demographic, clinical, laboratory, imaging, therapy and outcome data were extracted from electronic medical records. Clinical outcomes (discharges, mortality, length of stay) were followed up to February 20, 2020. The date of disease onset was defined as the day when the symptoms were noticed by patients.

### 2.3 Laboratory confirmation

Laboratory confirmation of SARS-CoV-2 was examined by two different institutions of Changsha or Loudi city simultaneously: the municipal Center for designated isolation hospitals and Disease Control and Prevention (CDC). RT-PCR detection reagents were provided by the two institutions. Throat swab specimens obtained from all patients at admission, were maintained in viral transport medium. SARS-CoV-2 was confirmed by RT-PCR using the same protocol described previously.^6^ To evaluate the overall lung involvement in CT imaging, each lung was divided into three lung zones, and each lung zone was assigned a score according to the percent of involvement.^7^

### 2.4 Clinical classification

Clinical classification was carried out with following standards:^5^ 1. mild type: mild clinical symptoms without pneumonia in chest imaging performance; 2. moderate type: fever, respiratory tract and other symptoms with pneumonia in imaging performance; 3. severe type: any one condition of the followings: (1) respiratory distress, respiratory rate ≥ 30 times/min; (2) fingertip oxygen saturation ≤ 93% in resting state; (3) arterial oxygen partial pressure/fraction of inspired oxygen ≤300 mmHg; 4. critical type: any one condition of the followings: (1) respiratory failure need mechanical ventilation; (2) shock; (3) other organs failure need intensive care unit (ICU) monitoring and treatment.

### 2.5 Statistical analysis

Continuous variables were described as medians and interquartile ranges (IQR). Categorical variables were summarized as counts and percentages. We categorized those four clinical types to three groups as mild group, moderate group and severe/critical group for further statistical analysis. Differences observed among all the three groups were analyzed by Chi-squared test or Fisher exact test for categorical data, one-way ANOVA or non-parametric Kruskal–Wallis test for quantitative data, as appropriate. Two-tailed p < 0.05 was thought as having significantly difference between three groups. For further pairwise comparisons, the significance level was adjusted using the Bonferroni method. All data were analyzed by SPSS (version 20.0; SPSS Inc., Chicago, IL, USA). Kaplan-Meier curve was constructed by the survival and survminer package in R, version 3.6.0 (http://www.r-project.org/).

## 3. Results

### 3.1 Epidemiological and baseline characteristics of patients

291 patients with laboratory-confirmed SARS-CoV-2 infection were included in this study. Epidemiological and baseline characteristics of patients in detail were shown in Table 1. For all the 291 patients, the median age was 46.0 years (IQR, 34.0 - 59.0 years; range, 1.0 - 84.0 years). Half patients (156, 53,6%) aged between 15 - 49 years, and 11 (3.8%) patients were aged below 15 years. 145 (49.8%) patients were male. 252 (86.6%) patients had a history of exposure to source of transmission within 14 days before symptoms onset. 114 (39.2%) patients were associated with familial clusters. 93 (32.0%) patients had at least one underlying disease including hypertension (13.4%), diabetes (7.6%), chronic liver disease (5.2%), cardiovascular disease (4.1%), chronic respiratory disease (3.4%), etc. The most common symptoms including fever (68.7%), cough (60.5%) and fatigue (31.6%). The median time from disease onset to first admission was 5 days (IQR: 3-8). None of the patients were medical staff. At admission, 245 (83.2%) patients had normal temperature (under 37.5°C), and the median highest body temperature during hospitalization was 38.0°C (37.2°C, 38.5°C).

**Table 1.** Demographics and baseline characteristics of 291 patients with COVID-19 in Hunan, China.

| Variables | All patients<br>(N=291) | Mild group (N=29) | Moderate group<br>(N=212) | Severe/critical<br>group (N=50) | p-values<br>($<0.050$) | Mild vs<br>Moderate<br>($<0.017$) | Mild vs<br>Severe/<br>critical<br>($<0.017$) | Moderate<br>vs Severe/<br>critical<br>($<0.017$) |
| --- | --- | --- | --- | --- | --- | --- | --- | --- |
| Age, years |  |  |  |  | 0.000 | 0.403 | 0.001 | 0.000 |
| Range | (1.0, 84.0) | (2.0, 76.0) | (2.0, 84.0) | (1.0, 82.0) |  |  |  |  |
| Medium (interquartile) | 46.0 (34.0, 59.0) | 50.0 (34.5, 53.0) | 41.0 (32.0, 53.0) | 60.5 (46.0, 70.0) |  |  |  |  |
| Age groups (years): |  |  |  |  | 0.000 |  |  |  |
| 0-14 | 11 (3.8%) | 2 (6.9%) | 8 (3.8%) | 1 (2.0%) |  |  |  |  |
| 15-49 | 156 (53.6%) | 12 (41.4%) | 131 (61.8%) | 13 (26.0%) |  |  |  |  |
| 50-64 | 80 (27.5%) | 13 (44.8%) | 51 (24.1%) | 16 (32.0%) |  |  |  |  |
| $\geq 65$ | 44 (15.1%) | 2 (6.9%) | 22 (10.4%) | 20 (40.0%) | | | | |
| Sex |  |  |  |  | 0.629 |  |  |  |
| Male | 145 (49.8%) | 16 (55.2%) | 102 (48.1%) | 27 (54.0%) |  |  |  |  |
| Female | 146 (50.2%) | 13 (44.8%) | 110 (51.9%) | 23 (46.0%) |  |  |  |  |
| Exposure to source of transmission within 14 days |  |  |  |  |  |  |  |  |
| Resident in Hubei | 70 (24.1%) | 6 (20.7%) | 53 (25.0%) | 11 (22.0%) | 0.819 |  |  |  |
| Recently been to Wuhan | 121 (41.6%) | 5 (17.2%) | 92 (43.4%) | 24 (48.0%) | 0.017 | 0.007 | 0.006 | 0.556 |
| Contacted with people from Wuhan | 41 (14.1%) | 4 (13.8%) | 27 (12.7%) | 10 (20.0%) | 0.414 |  |  |  |
| Contacted with local diagnosed patients | 112 (38.5%) | 14 (48.3%) | 83 (39.2%) | 15 (30.0%) | 0.255 |  |  |  |
| Any Expose History | 252 (86.6%) | 19 (65.5%) | 192 (90.6%) | 41 (82.0%) | 0.001 | 0.000 | 0.098 | 0.082 |
| Familial cluster | 114 (39.2%) | 16 (55.2%) | 82 (38.7%) | 16 (32.0%) | 0.121 |  |  |  |
| Coexisting conditions |  |  |  |  |  |  |  |  |
| Any | 93 (32.0%) | 10 (34.5%) | 54 (25.5%) | 29 (58.0%) | 0.000 | 0.303 | 0.044 | 0.000 |
| Chronic Respiratory Disease | 10 (3.4%) | 3 (10.3%) | 2 (0.9%) | 5 (10.0%) | 0.001 | 0.013 | 1.000 | 0.002 |
| Diabetes | 22 (7.6%) | 3 (10.3%) | 12 (5.7%) | 7 (14.0%) | 0.079 |  |  |  |
| Cardiovascular disease | 12 (4.1%) | 1 (3.4%) | 6 (2.8%) | 5 (10.0%) | 0.053 |  |  |  |
| Cerebrovascular diseases | 8 (2.7%) | 0 (0.0%) | 5 (2.4%) | 3 (6.0%) | 0.252 |  |  |  |
| Hypertension | 39 (13.4%) | 3 (10.3%) | 17 (8.0%) | 19 (38.0%) | 0.000 | 0.947 | 0.008 | 0.000 |
| Malignancy | 2 (0.7%) | 0 (0.0%) | 2 (0.9%) | 0 (0.0%) | 1.000 |  |  |  |
| Chronic liver disease | 15 (5.2%) | 4 (13.8%) | 9 (4.2%) | 2 (4.0%) | 0.100 |  |  |  |
| Chronic renal disease | 2 (0.7%) | 0 (0.0%) | 1 (0.5%) | 1 (2.0%) | 0.470 |  |  |  |
| Others | 23 (7.9%) | 0 (0.0%) | 17 (8.0%) | 6 (12.0%) | 0.159 |  |  |  |
| Signs and symptoms |  |  |  |  |  |  |  |  |
| Fever | 200 (68.7%) | 22 (75.9%) | 136 (64.2%) | 42 (84.0%) | 0.017 | 0.213 | 0.374 | 0.007 |
| Cough | 176 (60.5%) | 21 (72.4%) | 119 (56.1%) | 36 (72.0%) | 0.045 | 0.096 | 0.968 | 0.040 |
| Dyspnea | 37 (12.7%) | 3 (10.3%) | 16 (7.5%) | 18 (36.0%) | 0.000 | 0.875 | 0.013 | 0.000 |
| Myalgia | 27 (9.3%) | 3 (10.3%) | 18 (8.5%) | 6 (12.0%) | 0.671 |  |  |  |
| Headache | 20 (6.9%) | 2 (6.9%) | 13 (6.1%) | 5 (10.0%) | 0.548 |  |  |  |
| Dizziness | 12 (4.1%) | 1 (3.4%) | 10 (4.7%) | 1 (2.0%) | 0.883 |  |  |  |
| Abdominal pain | 1 (0.3%) | 0 (0.0%) | 0 (0.0%) | 1 (2.0%) | 0.271 |  |  |  |
| Diarrhoea | 25 (8.6%) | 3 (10.3%) | 17 (8.0%) | 5 (10.0%) | 0.737 |  |  |  |
| Fatigue | 92 (31.6%) | 5 (17.2%) | 61 (28.8%) | 26 (52.0%) | 0.001 | 0.191 | 0.002 | 0.002 |
| Nausea or vomiting | 17 (5.8%) | 6 (20.7%) | 9 (4.2%) | 2 (4.0%) | 0.006 | 0.002 | 0.047 | 1.000 |
| Pharyngalgia | 32 (11.0%) | 3 (10.3%) | 26 (12.3%) | 3 (6.0%) | 0.441 |  |  |  |
| Rhinorrhea | 7 (2.4%) | 2 (6.9%) | 3 (1.4%) | 2 (4.0%) | 0.071 |  |  |  |
| More than one sign or symptom | 199 (68.4%) | 23 (79.3%) | 134 (63.2%) | 42 (84.0%) | 0.007 | 0.088 | 0.599 | 0.005 |
| Sign Number | 2 (1, 3) | 2 (2, 3) | 2 (1, 3) | 3 (2, 4) | 0.000 | 0.074 | 0.136 | 0.000 |
| Vital Signs |  |  |  |  |  |  |  |  |
| Body temperature at admission (°C) | 36.9 (36.5, 37.4) | 36.7 (36.5, 37) | 36.9 (36.5, 37.3) | 36.9 (36.5, 37.6) | 0.321 |  |  |  |
| < 37.5 | 242 (83.2%) | 23 (79.3%) | 182 (85.8%) | 37 (74.0%) |  |  |  |  |
| 37.5-38.0 | 25 (8.6%) | 0 (0%) | 17 (8%) | 8 (16.0%) |  |  |  |  |
| 38.1-39.0 | 20 (6.9%) | 4 (13.8%) | 11 (5.2%) | 5 (10.0%) |  |  |  |  |
| >39.0 | 1 (0.3%) | 0 (0%) | 1 (0.5%) | 0 (0.0%) |  |  |  |  |
| Highest body temperature during hospitalization (°C) | 38.0 (37.2, 38.5) | 38.0 (37.0, 38.7) | 37.8 (37.2, 38.3) | 38.3 (37.7, 38.9) | 0.001 | 0.742 | 0.090 | 0.000 |
| < 37.5 | 96 (33.0%) | 9 (31.0%) | 78 (36.8%) | 9 (18.0%) |  |  |  |  |
| 37.5-38.0 | 75 (25.8%) | 7 (24.1%) | 56 (26.4%) | 12 (24.0%) |  |  |  |  |
| 38.1-39.0 | 97 (33.3%) | 9 (31.0%) | 66 (31.1%) | 22 (44.0%) |  |  |  |  |
| >39.0 | 22 (7.6%) | 4 (13.8%) | 11 (5.2%) | 7 (14.0%) |  |  |  |  |
| Respiratory rate | 20 (18, 21) | 20 (20, 21) | 20 (18, 20) | 20 (20, 22) | 0.000 | 0.003 | 0.587 | 0.000 |
| Pulse rate | 88 (80, 98) | 88 (78, 100) | 88 (80, 97) | 92 (78, 101) | 0.605 |  |  |  |
| Systolic blood pressure | 121 (114, 132) | 128 (110, 135) | 120 (114, 132) | 124 (114, 134) | 0.633 |  |  |  |
| Diastolic blood pressure | 78 (70, 84) | 74 (61, 86) | 78 (71, 84) | 78 (70, 84) | 0.313 |  |  |  |
| Mean arterial pressure | 93 (85, 99) | 92 (78, 100) | 93 (87, 99) | 92 (84, 100) | 0.617 |  |  |  |
| SpO<sub>2</sub> | 97 (96, 98) | n=0 | 97 (96, 98) | 96 (95, 97) | 0.000 | / | / | 0.000 |
| Time from illness onset to first hospital admission | 5.0 (3.0, 8.0) | 6.0 (3.0, 11.0) | 5.0 (3.0, 8.0) | 6.0 (4.0, 9.0) | 0.123 |  |  |  |

Values are numbers (percentages) or Medium (IQR) unless stated otherwise. Percentages do not total up to 100% owing to missing data.

After admission, 291 patients were categorized into mild group (29, 10.0%), moderate group (212, 72.8%) and severe/critical group (50, 17.2%) respectively. The age distribution was different among three groups (P<0.001). Patients in severe/critical group were older compared with the mild group (60.5 vs 50.0, p=0.001) or moderate group (60.5 vs 41.0, p<0.001). There was no difference in gender among three groups. Interestingly, as for the exposure history, the proportion of patients that had been to Wuhan in severe/critical group (17.2% vs 48.0%, p=0.006) and moderate group (17.2% vs 43.4%, p=0.007) were higher than that of mild group. Similar results were found in the history of any kinds of exposure to source of transmission. For coexisting conditions at admission, the proportion of patients with hypertension in the severe/critical group were higher than mild group (30.8% vs 10.3%, p=0.008) and moderate group (30.8% vs 8.0%, p=0.000). The result was also similar for chronic respiratory disease. A higher proportion of patients in severe/critical group had symptoms like fever, dyspnea and fatigue, while other symptoms like nausea or vomiting were more common in mild group.

### 3.2 Laboratory examinations

All patients received imaging examination including chest radiography or computed tomography (CT) on admission. Typical manifestations in CT and chest X-ray plain film on admission were bilateral and unilateral ground glass opacity or consolidation (Figure 1). Among all patients, 115 patients performed CT within one week after admission, which showed the medium score of lung involvement in both the whole lung field and each separate lung field (upper, middle, lower) in severe/critical group was higher than moderate group (all p < 0.01).

**Figure 1.**
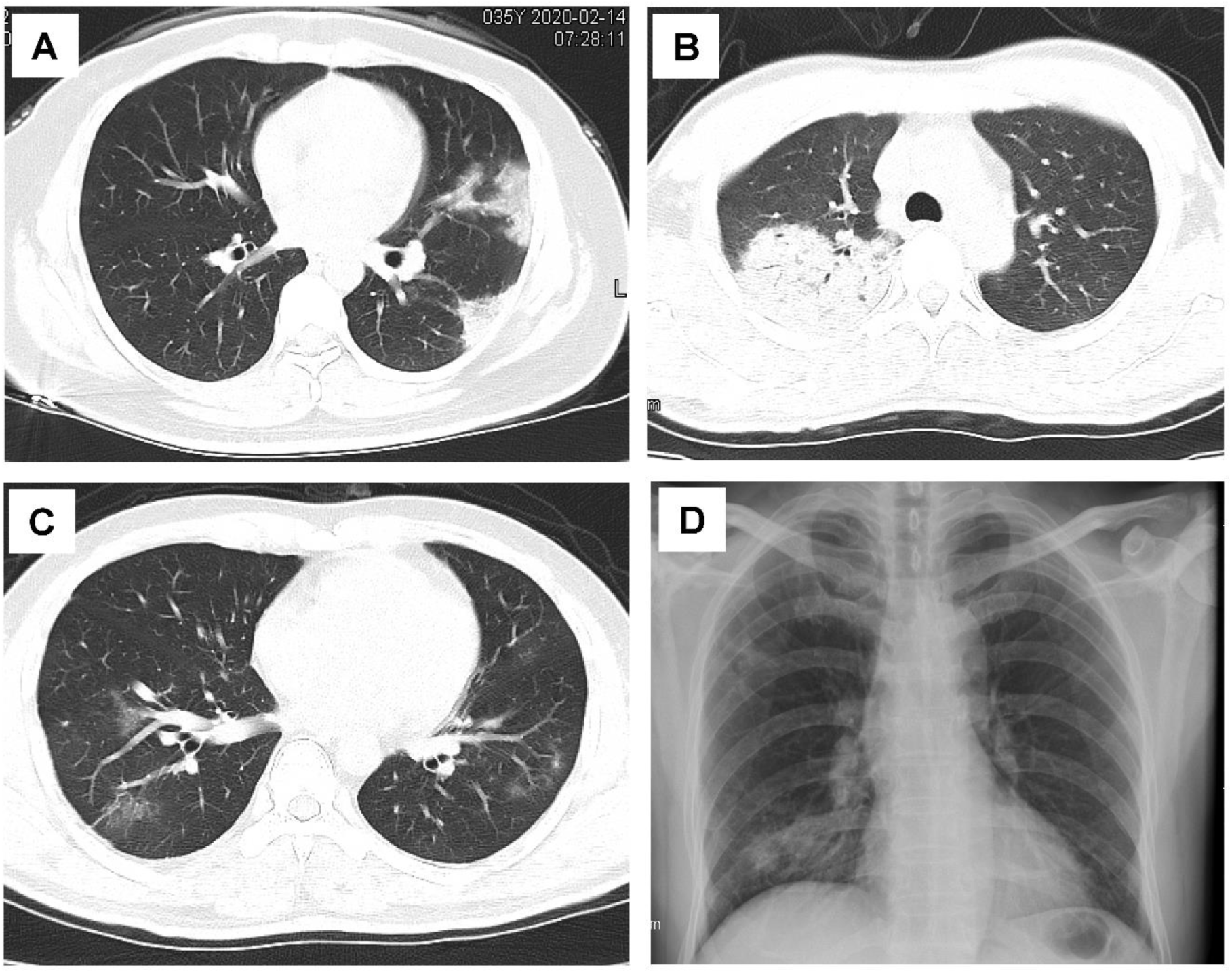
The typical manifestations in CT and chest X-ray for patients with COVID-19. Cross-sectional CT shows (A) patchy consolidation abutting the pleura in left upper lobe, (B) large areas of consolidation with air bronchogram sign in right upper lobe and (C) Ground glass opacity in bilateral upper lobe; (D) Chest X-ray shows patchy opacities of the right lower lung field.

Laboratory examinations data were given in Table 2. On admission, 36.1% or 22.7% patients showed leukopenia (white blood cell count < 4.00*10^9^/L) or lymphopenia (lymphocyte count < 0.8*10^9^/L) respectively. Neutrophils in 54 (18.6%) patients were below the normal range. Thrombocytopenia (platelet count <150*10^9^/L) were observed in 31.3% patients. Interestingly, 50.2% patients had eosinophils count lower than 0.02*10^9^/L, and the situation was obviously worse in severe/critical group whose medium eosinophils count was 0.00*10^9^/L. Compared with mild or moderate group, severe/critical group had a higher level of neutrophil and Neutrophil-to-Lymphocyte Ratio, and a lower level of lymphocytes, eosinophils and platelets (all p<0.01).

**Table 2.**
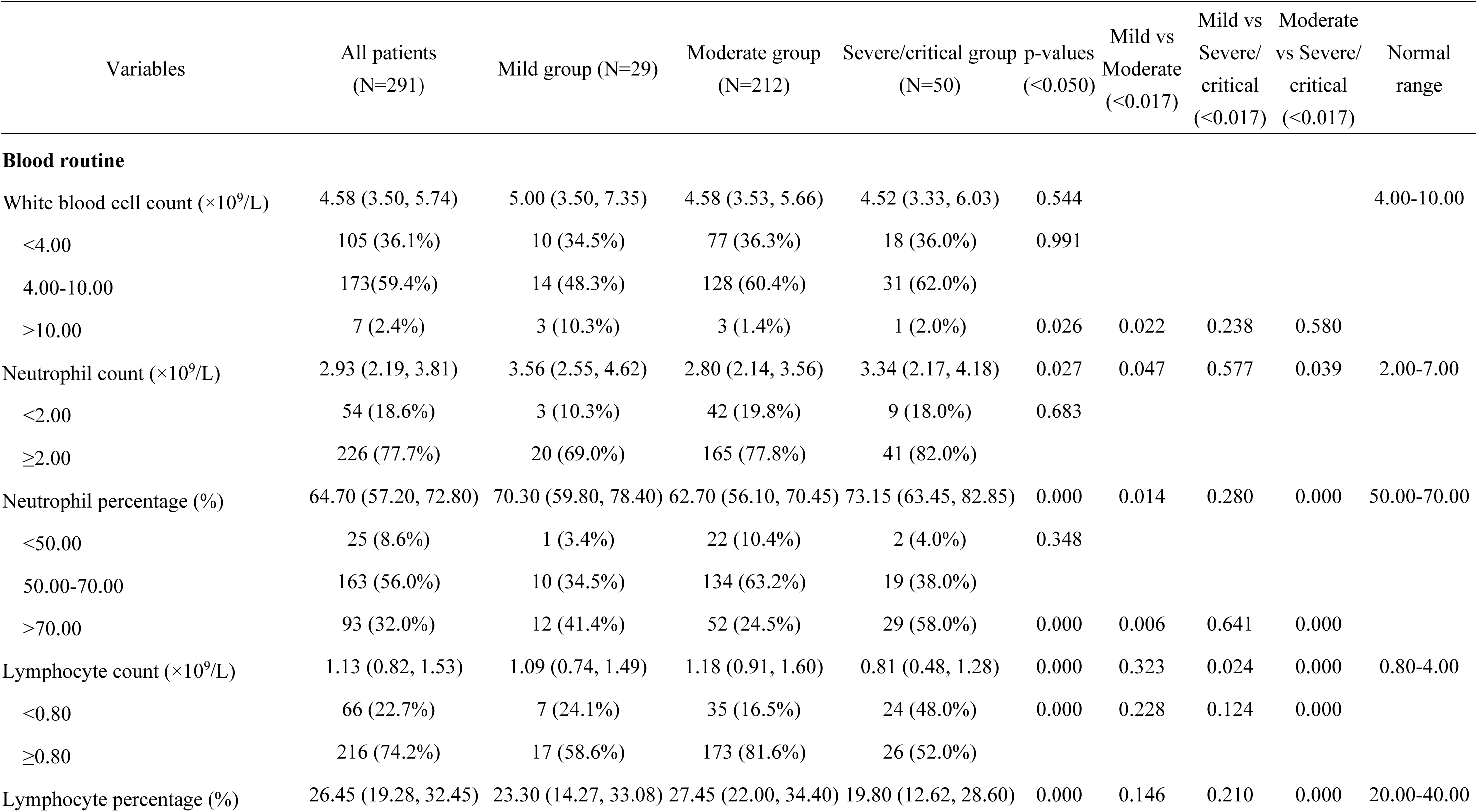
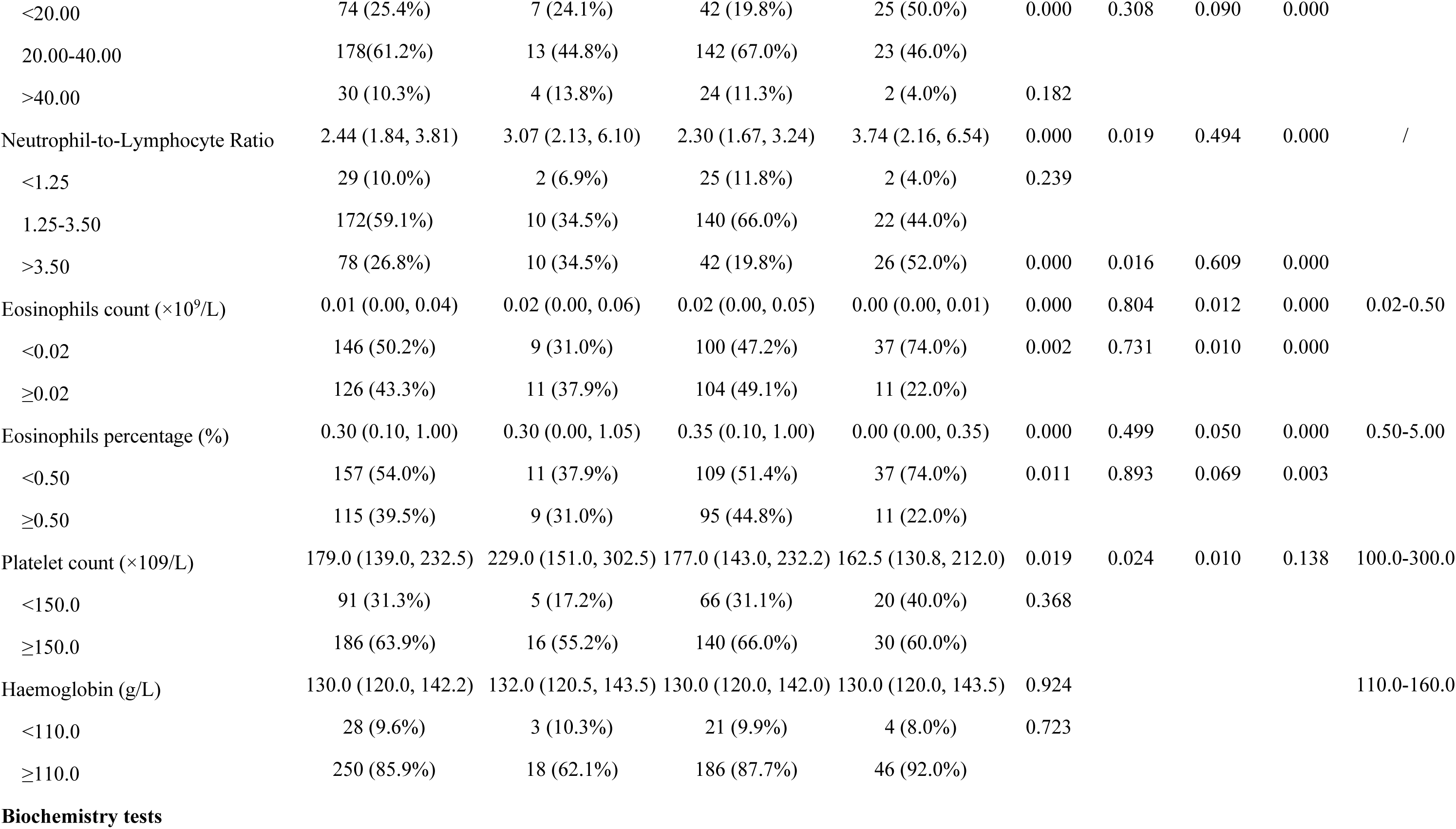
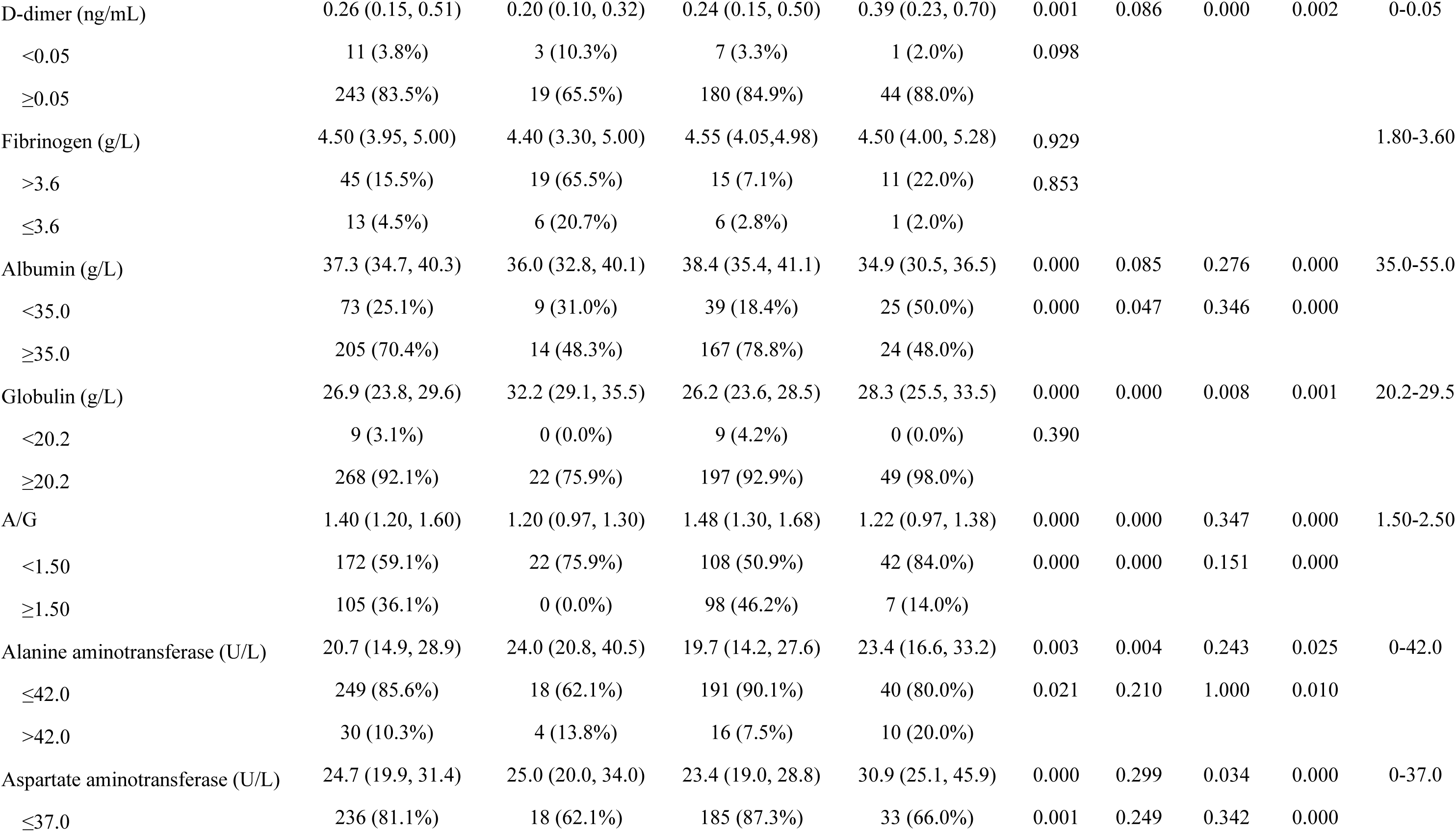
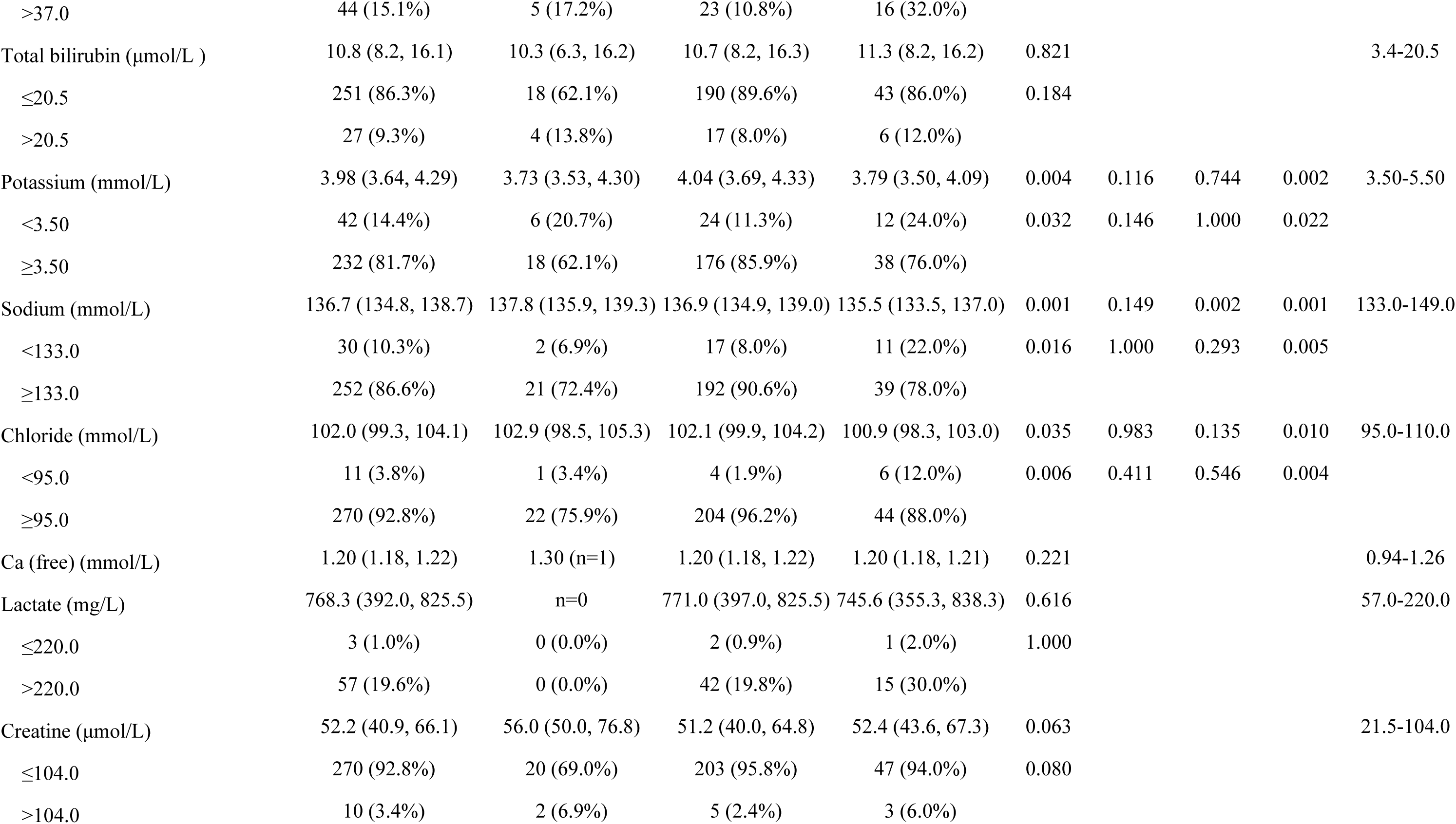
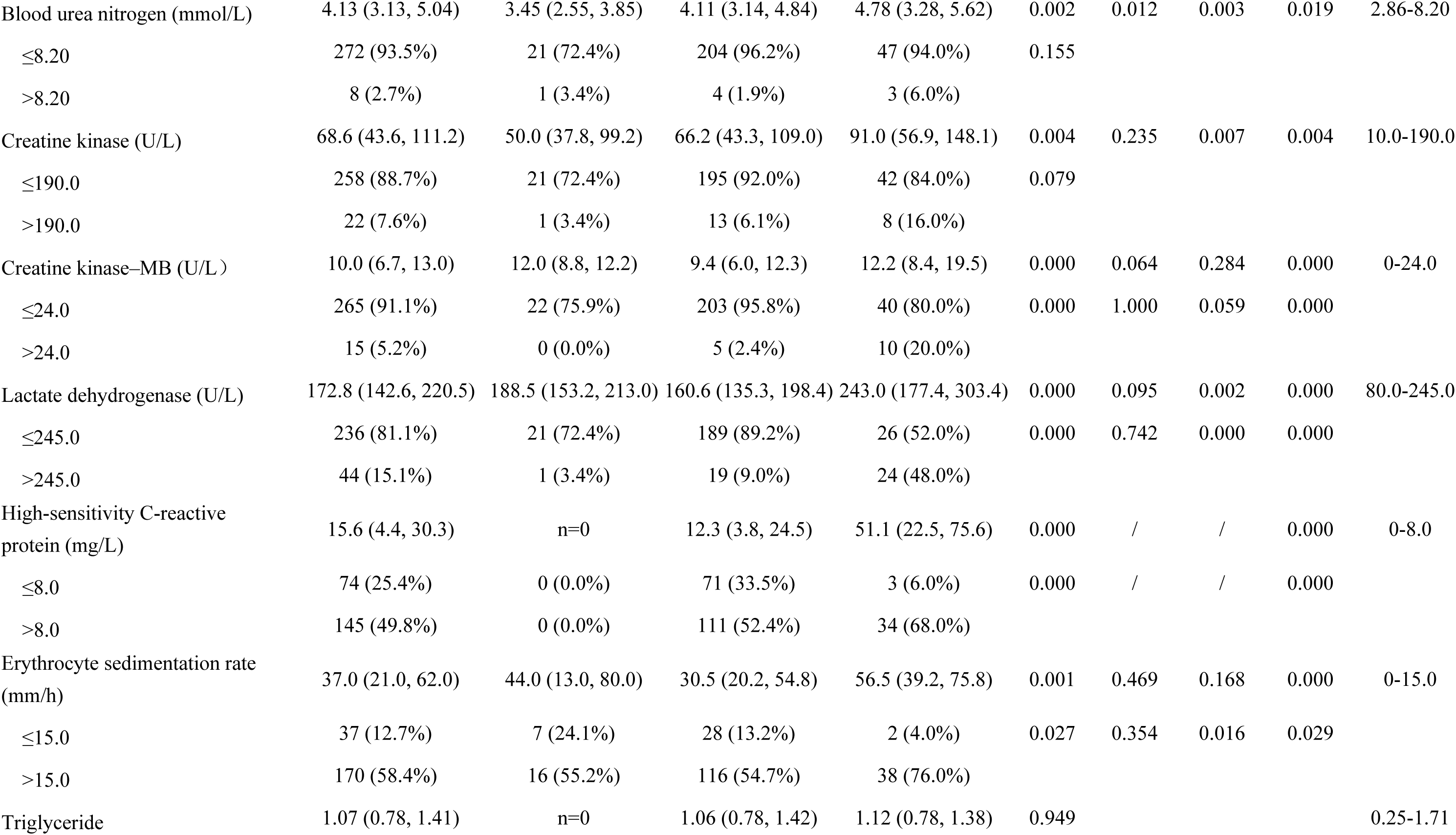
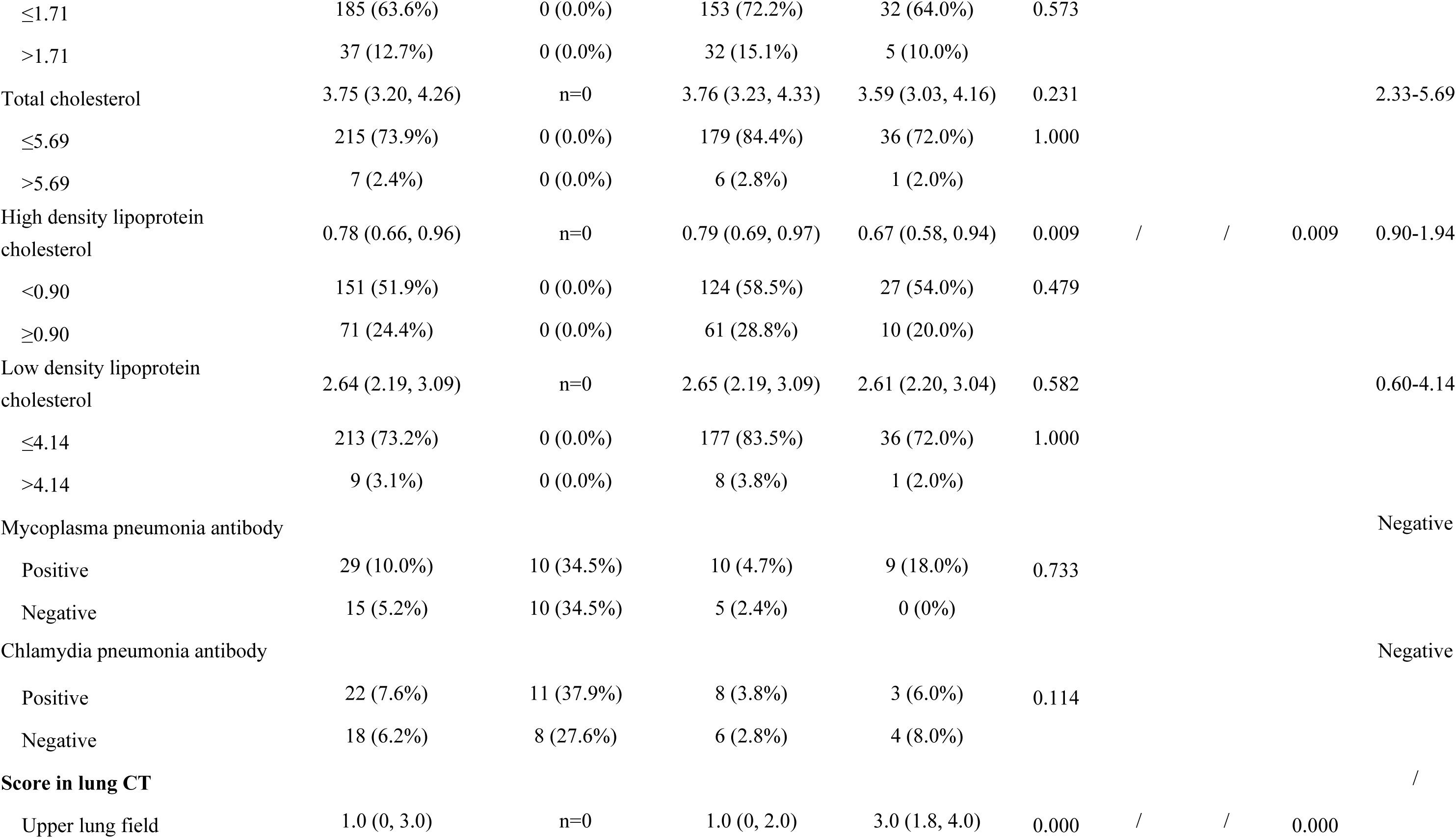
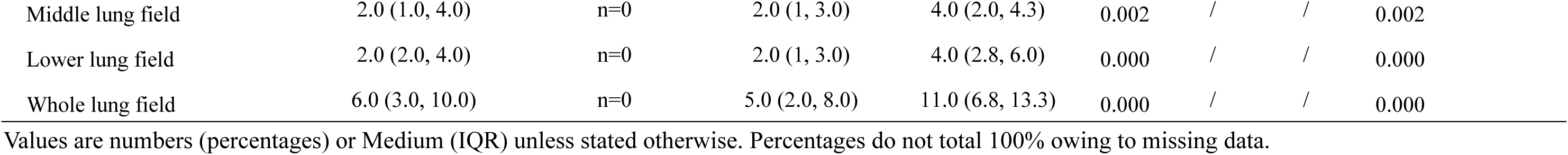
Laboratory findings of 291 patients with coronavirus disease 2019 (covid-19) in Hunan province, China.

| Variables | All patients<br>(N=291) | Mild group (N=29) | Moderate group<br>(N=212) | Severe/critical group<br>(N=50) | p-values<br>($<0.050$) | Mild vs<br>Moderate<br>($<0.017$) | Mild vs<br>Severe/<br>critical<br>($<0.017$) | Moderate<br>vs Severe/<br>critical<br>($<0.017$) | Normal<br>range |
| --- | --- | --- | --- | --- | --- | --- | --- | --- | --- |
| **Blood routine** |  |  |  |  |  |  |  |  |  |
| White blood cell count ($\times 10^9/L$) | 4.58 (3.50, 5.74) | 5.00 (3.50, 7.35) | 4.58 (3.53, 5.66) | 4.52 (3.33, 6.03) | 0.544 | | | | 4.00-10.00 |
| <4.00 | 105 (36.1%) | 10 (34.5%) | 77 (36.3%) | 18 (36.0%) | 0.991 |  |  |  |  |
| 4.00-10.00 | 173 (59.4%) | 14 (48.3%) | 128 (60.4%) | 31 (62.0%) |  |  |  |  |  |
| >10.00 | 7 (2.4%) | 3 (10.3%) | 3 (1.4%) | 1 (2.0%) | 0.026 | 0.022 | 0.238 | 0.580 |  |
| Neutrophil count ($\times 10^9/L$) | 2.93 (2.19, 3.81) | 3.56 (2.55, 4.62) | 2.80 (2.14, 3.56) | 3.34 (2.17, 4.18) | 0.027 | 0.047 | 0.577 | 0.039 | 2.00-7.00 |
| <2.00 | 54 (18.6%) | 3 (10.3%) | 42 (19.8%) | 9 (18.0%) | 0.683 |  |  |  |  |
| $\geq 2.00$ | 226 (77.7%) | 20 (69.0%) | 165 (77.8%) | 41 (82.0%) | | | | | |
| Neutrophil percentage (%) | 64.70 (57.20, 72.80) | 70.30 (59.80, 78.40) | 62.70 (56.10, 70.45) | 73.15 (63.45, 82.85) | 0.000 | 0.014 | 0.280 | 0.000 | 50.00-70.00 |
| <50.00 | 25 (8.6%) | 1 (3.4%) | 22 (10.4%) | 2 (4.0%) | 0.348 |  |  |  |  |
| 50.00-70.00 | 163 (56.0%) | 10 (34.5%) | 134 (63.2%) | 19 (38.0%) |  |  |  |  |  |
| >70.00 | 93 (32.0%) | 12 (41.4%) | 52 (24.5%) | 29 (58.0%) | 0.000 | 0.006 | 0.641 | 0.000 |  |
| Lymphocyte count ($\times 10^9/L$) | 1.13 (0.82, 1.53) | 1.09 (0.74, 1.49) | 1.18 (0.91, 1.60) | 0.81 (0.48, 1.28) | 0.000 | 0.323 | 0.024 | 0.000 | 0.80-4.00 |
| <0.80 | 66 (22.7%) | 7 (24.1%) | 35 (16.5%) | 24 (48.0%) | 0.000 | 0.228 | 0.124 | 0.000 |  |
| $\geq 0.80$ | 216 (74.2%) | 17 (58.6%) | 173 (81.6%) | 26 (52.0%) | | | | | |
| Lymphocyte percentage (%) | 26.45 (19.28, 32.45) | 23.30 (14.27, 33.08) | 27.45 (22.00, 34.40) | 19.80 (12.62, 28.60) | 0.000 | 0.146 | 0.210 | 0.000 | 20.00-40.00 |
| <20.00 | 74 (25.4%) | 7 (24.1%) | 42 (19.8%) | 25 (50.0%) | 0.000 | 0.308 | 0.090 | 0.000 |  |
| 20.00-40.00 | 178(61.2%) | 13 (44.8%) | 142 (67.0%) | 23 (46.0%) |  |  |  |  |  |
| >40.00 | 30 (10.3%) | 4 (13.8%) | 24 (11.3%) | 2 (4.0%) | 0.182 |  |  |  |  |
| Neutrophil-to-Lymphocyte Ratio | 2.44 (1.84, 3.81) | 3.07 (2.13, 6.10) | 2.30 (1.67, 3.24) | 3.74 (2.16, 6.54) | 0.000 | 0.019 | 0.494 | 0.000 | / |
| <1.25 | 29 (10.0%) | 2 (6.9%) | 25 (11.8%) | 2 (4.0%) | 0.239 |  |  |  |  |
| 1.25-3.50 | 172(59.1%) | 10 (34.5%) | 140 (66.0%) | 22 (44.0%) |  |  |  |  |  |
| >3.50 | 78 (26.8%) | 10 (34.5%) | 42 (19.8%) | 26 (52.0%) | 0.000 | 0.016 | 0.609 | 0.000 |  |
| Eosinophils count (×10<sup>9</sup>/L) | 0.01 (0.00, 0.04) | 0.02 (0.00, 0.06) | 0.02 (0.00, 0.05) | 0.00 (0.00, 0.01) | 0.000 | 0.804 | 0.012 | 0.000 | 0.02-0.50 |
| <0.02 | 146 (50.2%) | 9 (31.0%) | 100 (47.2%) | 37 (74.0%) | 0.002 | 0.731 | 0.010 | 0.000 |  |
| ≥0.02 | 126 (43.3%) | 11 (37.9%) | 104 (49.1%) | 11 (22.0%) |  |  |  |  |  |
| Eosinophils percentage (%) | 0.30 (0.10, 1.00) | 0.30 (0.00, 1.05) | 0.35 (0.10, 1.00) | 0.00 (0.00, 0.35) | 0.000 | 0.499 | 0.050 | 0.000 | 0.50-5.00 |
| <0.50 | 157 (54.0%) | 11 (37.9%) | 109 (51.4%) | 37 (74.0%) | 0.011 | 0.893 | 0.069 | 0.003 |  |
| ≥0.50 | 115 (39.5%) | 9 (31.0%) | 95 (44.8%) | 11 (22.0%) |  |  |  |  |  |
| Platelet count (×10<sup>9</sup>/L) | 179.0 (139.0, 232.5) | 229.0 (151.0, 302.5) | 177.0 (143.0, 232.2) | 162.5 (130.8, 212.0) | 0.019 | 0.024 | 0.010 | 0.138 | 100.0-300.0 |
| <150.0 | 91 (31.3%) | 5 (17.2%) | 66 (31.1%) | 20 (40.0%) | 0.368 |  |  |  |  |
| ≥150.0 | 186 (63.9%) | 16 (55.2%) | 140 (66.0%) | 30 (60.0%) |  |  |  |  |  |
| Haemoglobin (g/L) | 130.0 (120.0, 142.2) | 132.0 (120.5, 143.5) | 130.0 (120.0, 142.0) | 130.0 (120.0, 143.5) | 0.924 |  |  |  | 110.0-160.0 |
| <110.0 | 28 (9.6%) | 3 (10.3%) | 21 (9.9%) | 4 (8.0%) | 0.723 |  |  |  |  |
| ≥110.0 | 250 (85.9%) | 18 (62.1%) | 186 (87.7%) | 46 (92.0%) |  |  |  |  |  |
| Biochemistry tests |  |  |  |  |  |  |  |  |  |
| D-dimer (ng/mL) | 0.26 (0.15, 0.51) | 0.20 (0.10, 0.32) | 0.24 (0.15, 0.50) | 0.39 (0.23, 0.70) | 0.001 | 0.086 | 0.000 | 0.002 | 0-0.05 |
| <0.05 | 11 (3.8%) | 3 (10.3%) | 7 (3.3%) | 1 (2.0%) | 0.098 |  |  |  |  |
| ≥0.05 | 243 (83.5%) | 19 (65.5%) | 180 (84.9%) | 44 (88.0%) |  |  |  |  |  |
| Fibrinogen (g/L) | 4.50 (3.95, 5.00) | 4.40 (3.30, 5.00) | 4.55 (4.05,4.98) | 4.50 (4.00, 5.28) | 0.929 |  |  |  | 1.80-3.60 |
| >3.6 | 45 (15.5%) | 19 (65.5%) | 15 (7.1%) | 11 (22.0%) | 0.853 |  |  |  |  |
| ≤3.6 | 13 (4.5%) | 6 (20.7%) | 6 (2.8%) | 1 (2.0%) |  |  |  |  |  |
| Albumin (g/L) | 37.3 (34.7, 40.3) | 36.0 (32.8, 40.1) | 38.4 (35.4, 41.1) | 34.9 (30.5, 36.5) | 0.000 | 0.085 | 0.276 | 0.000 | 35.0-55.0 |
| <35.0 | 73 (25.1%) | 9 (31.0%) | 39 (18.4%) | 25 (50.0%) | 0.000 | 0.047 | 0.346 | 0.000 |  |
| ≥35.0 | 205 (70.4%) | 14 (48.3%) | 167 (78.8%) | 24 (48.0%) |  |  |  |  |  |
| Globulin (g/L) | 26.9 (23.8, 29.6) | 32.2 (29.1, 35.5) | 26.2 (23.6, 28.5) | 28.3 (25.5, 33.5) | 0.000 | 0.000 | 0.008 | 0.001 | 20.2-29.5 |
| <20.2 | 9 (3.1%) | 0 (0.0%) | 9 (4.2%) | 0 (0.0%) | 0.390 |  |  |  |  |
| ≥20.2 | 268 (92.1%) | 22 (75.9%) | 197 (92.9%) | 49 (98.0%) |  |  |  |  |  |
| A/G | 1.40 (1.20, 1.60) | 1.20 (0.97, 1.30) | 1.48 (1.30, 1.68) | 1.22 (0.97, 1.38) | 0.000 | 0.000 | 0.347 | 0.000 | 1.50-2.50 |
| <1.50 | 172 (59.1%) | 22 (75.9%) | 108 (50.9%) | 42 (84.0%) | 0.000 | 0.000 | 0.151 | 0.000 |  |
| ≥1.50 | 105 (36.1%) | 0 (0.0%) | 98 (46.2%) | 7 (14.0%) |  |  |  |  |  |
| Alanine aminotransferase (U/L) | 20.7 (14.9, 28.9) | 24.0 (20.8, 40.5) | 19.7 (14.2, 27.6) | 23.4 (16.6, 33.2) | 0.003 | 0.004 | 0.243 | 0.025 | 0-42.0 |
| ≤42.0 | 249 (85.6%) | 18 (62.1%) | 191 (90.1%) | 40 (80.0%) | 0.021 | 0.210 | 1.000 | 0.010 |  |
| >42.0 | 30 (10.3%) | 4 (13.8%) | 16 (7.5%) | 10 (20.0%) |  |  |  |  |  |
| Aspartate aminotransferase (U/L) | 24.7 (19.9, 31.4) | 25.0 (20.0, 34.0) | 23.4 (19.0, 28.8) | 30.9 (25.1, 45.9) | 0.000 | 0.299 | 0.034 | 0.000 | 0-37.0 |
| ≤37.0 | 236 (81.1%) | 18 (62.1%) | 185 (87.3%) | 33 (66.0%) | 0.001 | 0.249 | 0.342 | 0.000 |  |
| >37.0 | 44 (15.1%) | 5 (17.2%) | 23 (10.8%) | 16 (32.0%) |  |  |  |  |  |
| Total bilirubin (μmol/L ) | 10.8 (8.2, 16.1) | 10.3 (6.3, 16.2) | 10.7 (8.2, 16.3) | 11.3 (8.2, 16.2) | 0.821 |  |  |  | 3.4-20.5 |
| ≤20.5 | 251 (86.3%) | 18 (62.1%) | 190 (89.6%) | 43 (86.0%) | 0.184 |  |  |  |  |
| >20.5 | 27 (9.3%) | 4 (13.8%) | 17 (8.0%) | 6 (12.0%) |  |  |  |  |  |
| Potassium (mmol/L) | 3.98 (3.64, 4.29) | 3.73 (3.53, 4.30) | 4.04 (3.69, 4.33) | 3.79 (3.50, 4.09) | 0.004 | 0.116 | 0.744 | 0.002 | 3.50-5.50 |
| <3.50 | 42 (14.4%) | 6 (20.7%) | 24 (11.3%) | 12 (24.0%) | 0.032 | 0.146 | 1.000 | 0.022 |  |
| ≥3.50 | 232 (81.7%) | 18 (62.1%) | 176 (85.9%) | 38 (76.0%) |  |  |  |  |  |
| Sodium (mmol/L) | 136.7 (134.8, 138.7) | 137.8 (135.9, 139.3) | 136.9 (134.9, 139.0) | 135.5 (133.5, 137.0) | 0.001 | 0.149 | 0.002 | 0.001 | 133.0-149.0 |
| <133.0 | 30 (10.3%) | 2 (6.9%) | 17 (8.0%) | 11 (22.0%) | 0.016 | 1.000 | 0.293 | 0.005 |  |
| ≥133.0 | 252 (86.6%) | 21 (72.4%) | 192 (90.6%) | 39 (78.0%) |  |  |  |  |  |
| Chloride (mmol/L) | 102.0 (99.3, 104.1) | 102.9 (98.5, 105.3) | 102.1 (99.9, 104.2) | 100.9 (98.3, 103.0) | 0.035 | 0.983 | 0.135 | 0.010 | 95.0-110.0 |
| <95.0 | 11 (3.8%) | 1 (3.4%) | 4 (1.9%) | 6 (12.0%) | 0.006 | 0.411 | 0.546 | 0.004 |  |
| ≥95.0 | 270 (92.8%) | 22 (75.9%) | 204 (96.2%) | 44 (88.0%) |  |  |  |  |  |
| Ca (free) (mmol/L) | 1.20 (1.18, 1.22) | 1.30 (n=1) | 1.20 (1.18, 1.22) | 1.20 (1.18, 1.21) | 0.221 |  |  |  | 0.94-1.26 |
| Lactate (mg/L) | 768.3 (392.0, 825.5) | n=0 | 771.0 (397.0, 825.5) | 745.6 (355.3, 838.3) | 0.616 |  |  |  | 57.0-220.0 |
| ≤220.0 | 3 (1.0%) | 0 (0.0%) | 2 (0.9%) | 1 (2.0%) | 1.000 |  |  |  |  |
| >220.0 | 57 (19.6%) | 0 (0.0%) | 42 (19.8%) | 15 (30.0%) |  |  |  |  |  |
| Creatine (μmol/L) | 52.2 (40.9, 66.1) | 56.0 (50.0, 76.8) | 51.2 (40.0, 64.8) | 52.4 (43.6, 67.3) | 0.063 |  |  |  | 21.5-104.0 |
| ≤104.0 | 270 (92.8%) | 20 (69.0%) | 203 (95.8%) | 47 (94.0%) | 0.080 |  |  |  |  |
| >104.0 | 10 (3.4%) | 2 (6.9%) | 5 (2.4%) | 3 (6.0%) |  |  |  |  |  |
| Blood urea nitrogen (mmol/L) | 4.13 (3.13, 5.04) | 3.45 (2.55, 3.85) | 4.11 (3.14, 4.84) | 4.78 (3.28, 5.62) | 0.002 | 0.012 | 0.003 | 0.019 | 2.86-8.20 |
| ≤8.20 | 272 (93.5%) | 21 (72.4%) | 204 (96.2%) | 47 (94.0%) | 0.155 |  |  |  |  |
| >8.20 | 8 (2.7%) | 1 (3.4%) | 4 (1.9%) | 3 (6.0%) |  |  |  |  |  |
| Creatine kinase (U/L) | 68.6 (43.6, 111.2) | 50.0 (37.8, 99.2) | 66.2 (43.3, 109.0) | 91.0 (56.9, 148.1) | 0.004 | 0.235 | 0.007 | 0.004 | 10.0-190.0 |
| ≤190.0 | 258 (88.7%) | 21 (72.4%) | 195 (92.0%) | 42 (84.0%) | 0.079 |  |  |  |  |
| >190.0 | 22 (7.6%) | 1 (3.4%) | 13 (6.1%) | 8 (16.0%) |  |  |  |  |  |
| Creatine kinase-MB (U/L) | 10.0 (6.7, 13.0) | 12.0 (8.8, 12.2) | 9.4 (6.0, 12.3) | 12.2 (8.4, 19.5) | 0.000 | 0.064 | 0.284 | 0.000 | 0-24.0 |
| ≤24.0 | 265 (91.1%) | 22 (75.9%) | 203 (95.8%) | 40 (80.0%) | 0.000 | 1.000 | 0.059 | 0.000 |  |
| >24.0 | 15 (5.2%) | 0 (0.0%) | 5 (2.4%) | 10 (20.0%) |  |  |  |  |  |
| Lactate dehydrogenase (U/L) | 172.8 (142.6, 220.5) | 188.5 (153.2, 213.0) | 160.6 (135.3, 198.4) | 243.0 (177.4, 303.4) | 0.000 | 0.095 | 0.002 | 0.000 | 80.0-245.0 |
| ≤245.0 | 236 (81.1%) | 21 (72.4%) | 189 (89.2%) | 26 (52.0%) | 0.000 | 0.742 | 0.000 | 0.000 |  |
| >245.0 | 44 (15.1%) | 1 (3.4%) | 19 (9.0%) | 24 (48.0%) |  |  |  |  |  |
| High-sensitivity C-reactive protein (mg/L) | 15.6 (4.4, 30.3) | n=0 | 12.3 (3.8, 24.5) | 51.1 (22.5, 75.6) | 0.000 | / | / | 0.000 | 0-8.0 |
| ≤8.0 | 74 (25.4%) | 0 (0.0%) | 71 (33.5%) | 3 (6.0%) | 0.000 | / | / | 0.000 |  |
| >8.0 | 145 (49.8%) | 0 (0.0%) | 111 (52.4%) | 34 (68.0%) |  |  |  |  |  |
| Erythrocyte sedimentation rate (mm/h) | 37.0 (21.0, 62.0) | 44.0 (13.0, 80.0) | 30.5 (20.2, 54.8) | 56.5 (39.2, 75.8) | 0.001 | 0.469 | 0.168 | 0.000 | 0-15.0 |
| ≤15.0 | 37 (12.7%) | 7 (24.1%) | 28 (13.2%) | 2 (4.0%) | 0.027 | 0.354 | 0.016 | 0.029 |  |
| >15.0 | 170 (58.4%) | 16 (55.2%) | 116 (54.7%) | 38 (76.0%) |  |  |  |  |  |
| Triglyceride | 1.07 (0.78, 1.41) | n=0 | 1.06 (0.78, 1.42) | 1.12 (0.78, 1.38) | 0.949 |  |  |  | 0.25-1.71 |
| ≤1.71 | 185 (63.6%) | 0 (0.0%) | 153 (72.2%) | 32 (64.0%) | 0.573 |  |  |  |  |
| >1.71 | 37 (12.7%) | 0 (0.0%) | 32 (15.1%) | 5 (10.0%) |  |  |  |  |  |
| Total cholesterol | 3.75 (3.20, 4.26) | n=0 | 3.76 (3.23, 4.33) | 3.59 (3.03, 4.16) | 0.231 |  |  |  | 2.33-5.69 |
| ≤5.69 | 215 (73.9%) | 0 (0.0%) | 179 (84.4%) | 36 (72.0%) | 1.000 |  |  |  |  |
| >5.69 | 7 (2.4%) | 0 (0.0%) | 6 (2.8%) | 1 (2.0%) |  |  |  |  |  |
| High density lipoprotein cholesterol | 0.78 (0.66, 0.96) | n=0 | 0.79 (0.69, 0.97) | 0.67 (0.58, 0.94) | 0.009 | / | / | 0.009 | 0.90-1.94 |
| <0.90 | 151 (51.9%) | 0 (0.0%) | 124 (58.5%) | 27 (54.0%) | 0.479 |  |  |  |  |
| ≥0.90 | 71 (24.4%) | 0 (0.0%) | 61 (28.8%) | 10 (20.0%) |  |  |  |  |  |
| Low density lipoprotein cholesterol | 2.64 (2.19, 3.09) | n=0 | 2.65 (2.19, 3.09) | 2.61 (2.20, 3.04) | 0.582 |  |  |  | 0.60-4.14 |
| ≤4.14 | 213 (73.2%) | 0 (0.0%) | 177 (83.5%) | 36 (72.0%) | 1.000 |  |  |  |  |
| >4.14 | 9 (3.1%) | 0 (0.0%) | 8 (3.8%) | 1 (2.0%) |  |  |  |  |  |
| Mycoplasma pneumonia antibody |  |  |  |  |  |  |  |  | Negative |
| Positive | 29 (10.0%) | 10 (34.5%) | 10 (4.7%) | 9 (18.0%) | 0.733 |  |  |  |  |
| Negative | 15 (5.2%) | 10 (34.5%) | 5 (2.4%) | 0 (0%) |  |  |  |  |  |
| Chlamydia pneumonia antibody |  |  |  |  |  |  |  |  | Negative |
| Positive | 22 (7.6%) | 11 (37.9%) | 8 (3.8%) | 3 (6.0%) | 0.114 |  |  |  |  |
| Negative | 18 (6.2%) | 8 (27.6%) | 6 (2.8%) | 4 (8.0%) |  |  |  |  |  |
| **Score in lung CT** |  |  |  |  |  |  |  |  | / |
| Upper lung field | 1.0 (0, 3.0) | n=0 | 1.0 (0, 2.0) | 3.0 (1.8, 4.0) | 0.000 | / | / | 0.000 |  |

|  |  |  |  |  |  |  |  |  |
| --- | --- | --- | --- | --- | --- | --- | --- | --- |
| Middle lung field | 2.0 (1.0, 4.0) | n=0 | 2.0 (1, 3.0) | 4.0 (2.0, 4.3) | 0.002 | / | / | 0.002 |
| Lower lung field | 2.0 (2.0, 4.0) | n=0 | 2.0 (1, 3.0) | 4.0 (2.8, 6.0) | 0.000 | / | / | 0.000 |
| Whole lung field | 6.0 (3.0, 10.0) | n=0 | 5.0 (2.0, 8.0) | 11.0 (6.8, 13.3) | 0.000 | / | / | 0.000 |

Values are numbers (percentages) or Medium (IQR) unless stated otherwise. Percentages do not total 100% owing to missing data.

Biochemistry tests showed 83.5% patients had an increased level of D-dimer and the level was higher in severe/critical group than in mild group (0.39 vs 0.20, p=0.000) and moderate group (0.39 vs 0.24, p=0.002). The coagulation function test showed an increase of fibrinogen in 45 of 58 (77.6%) patients with available results. For liver function test, severe/critical group had a higher proportion of elevated aspartate aminotransferase (AST) than mild group (32.0% vs 17.2%, p<0.001). No obvious abnormality of renal function was found in patients without any history of kidney disease. 10.3% patients had hyponatremia and the proportion were higher in severe/critical group than in mild group (22.0% vs 6.9%, p=0.005). Moreover, erythrocyte sedimentation rate (ESR) and high-sensitivity C-reactive protein (h-CRP) were increased in more than 50% of patients. Decreased level of high-density lipoprotein cholesterol (HDL) was detected in 51.9% patients. Compared with mild or moderate group, severe/critical group had a higher level of h-CRP, ESR, creatine kinase, creatine kinase–MB, lactate dehydrogenase, and lower level of sodium and HDL (all p<0.01). In patients with available Mycoplasma pneumonia or Chlamydia pneumonia antibody test, 29 of 44 (65.9%) or 22 of 40 (55.0%) patients had positive antibody test results respectively.

### 3.3 Treatment and prognosis

285 (97.9%) of 291 patients received antiviral therapy (Table 3), including lopinavir and ritonavir tablets (75.9%), recombinant human interferon α2b (45.4%), recombinant cytokine gene derived protein (18.9%) and arbidol hydrochloride capsules (17.2%). 281 (96.6%) patients were treated with Chinese Medicine. The dose and method of administration was based on the guidelines^5^.

**Table 3.** Treatments and outcomes of 291 patients with COVID-19 in Hunan, China.

| Variables | All patients(N=291) | Mild group<br>(N=29) | Moderate group<br>(N=212) | Severe/critical<br>group (N=50) | p-values<br>(<0.050) | Mild vs<br>Moderate<br>(<0.017) | Mild vs<br>Severe/<br>critical<br>(<0.017) | Moderate<br>vs Severe/<br>critical<br>(<0.017) |
| --- | --- | --- | --- | --- | --- | --- | --- | --- |
| Antivirus therapy |  |  |  |  |  |  |  |  |
| Any | 285 (97.9%) | 28 (96.6%) | 207 (97.6%) | 50 (100%) | 0.368 |  |  |  |
| Lopinavir and ritonavir tablets | 221 (75.9%) | 27 (93.1%) | 153 (72.2%) | 41 (82.0%) | 0.026 | 0.015 | 0.300 | 0.154 |
| Recombinant human interferon α2b | 132 (45.4%) | 24 (82.8%) | 91 (42.9%) | 17 (34.0%) | 0.000 | 0.000 | 0.000 | 0.249 |
| Recombinant Cytokine Gene Derived Protein | 55 (18.9%) | 0 (0.0%) | 52 (24.5%) | 3 (6.0%) | 0.000 | 0.003 | 0.463 | 0.004 |
| Arbidol Hydrochloride Capsules | 50 (17.2%) | 14 (48.3%) | 27 (12.7%) | 9 (18.0%) | 0.000 | 0.000 | 0.004 | 0.331 |
| Oseltamivir | 1 (0.3%) | 0 (0.0%) | 0 (0.0%) | 1 (2.0%) | 0.271 |  |  |  |
| Chloroquine | 2 (0.7%) | 0 (0.0%) | 2 (0.9%) | 0 (0.0%) | 1.000 |  |  |  |
| Combined | 157 (54.0%) | 7 (24.1%) | 127 (59.9%) | 23 (46.0%) | 0.001 | 0.000 | 0.054 | 0.074 |
| Chinese Medicine | 281 (96.6%) | 26 (89.7%) | 208 (98.1%) | 47 (94.0%) | 0.027 | 0.039 | 0.793 | 0.256 |
| Glucocorticoid | 67 (23%) | 13 (44.8%) | 46 (21.7%) | 8 (16%) | 0.009 | 0.007 | 0.005 | 0.370 |
| Length of stay |  |  |  |  |  |  |  |  |
| Discharged patients | 12.0 (10.0, 15.0) | 16.0 (12.5, 21.5) | 12.0 (9.0, 14.0) | 14.0 (13.0, 17.0) | 0.000 | 0.015 | 0.370 | 0.000 |
| Outcomes |  |  |  |  | 0.036 |  |  |  |
| Death | 2 (0.7%) | 0 (0.0%) | 1 (0.5%) | 1 (2.0%) |  |  |  |  |
| Remained in hospital | 130 (44.7%) | 20 (69.0%) | 90 (42.5%) | 20 (40.0%) |  |  |  |  |
| Discharge | 159 (54.6%) | 9 (31.0%) | 121 (57.1%) | 29 (58.0%) |  |  |  |  |

Values are numbers (percentages) or Medium (IQR) unless stated otherwise.

Patients were discharged from hospital when the symptoms relieved, chest imaging improved, fever abated for at least three days, and two samples taken from respiratory tract 1 day apart were negative for SARS-Cov-2 RNA. Until February 20, 2020, 159(54.6%) of all patients had been discharged. The length of hospitalization ranged from 5 to 25 days in discharged patients. We used a Kaplan-Meier plot to analyze the length of hospitalization for all 291 patients (Figure 2), which shows a median length of 16 days (IQR 14-17). 2 of 291 (0.7%) patients died during hospitalization, both of whom had an exposure history of having been to Wuhan within 14 days prior to onset of illness.

**Figure 2.**
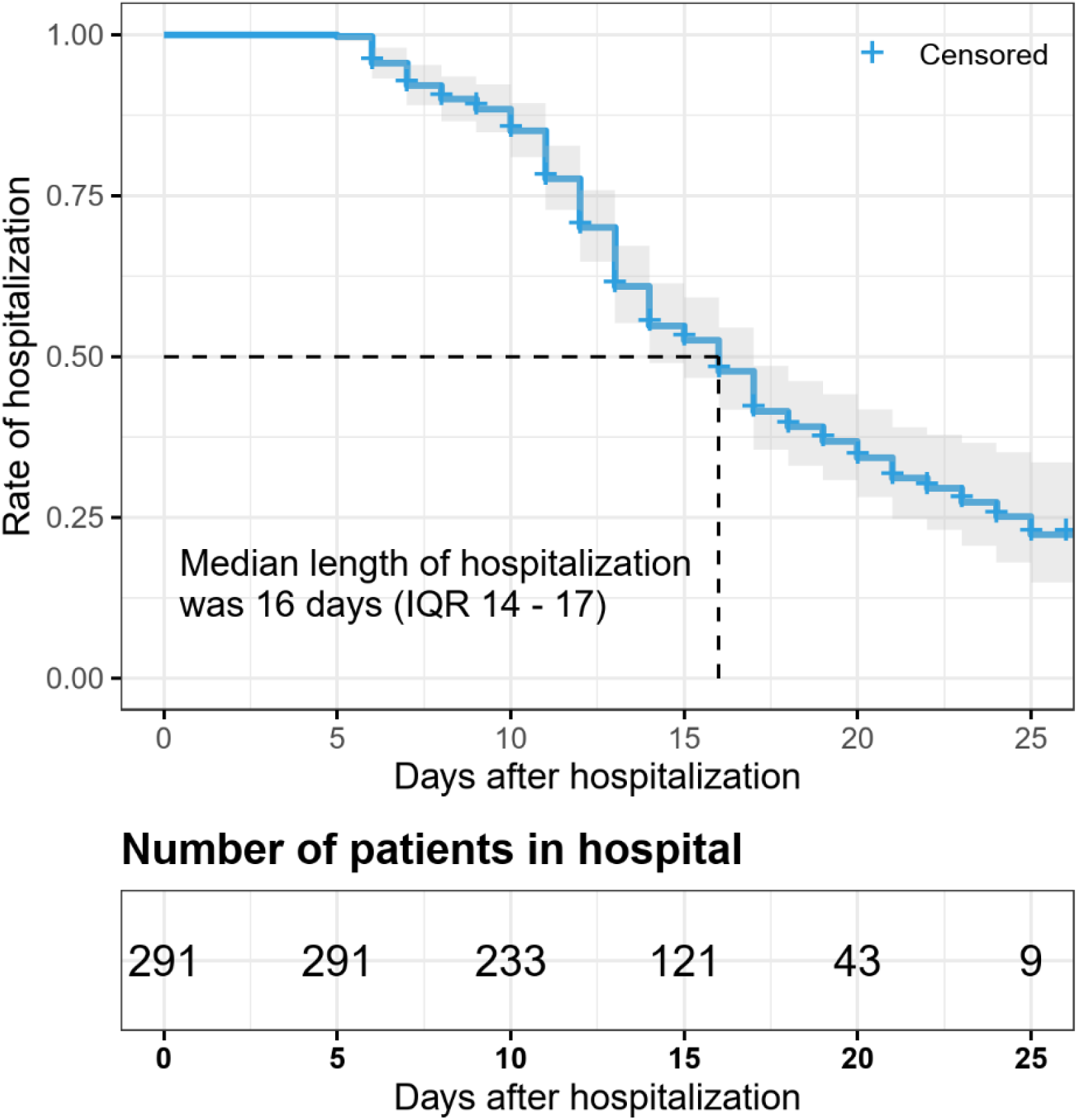
Kaplan–Meier analysis of the length of hospitalization for patients with COVID-19. Shaded areas represent the 95% confidence intervals based on Kaplan-Meier estimation. The median length of hospitalization for all patients was 16 days (IQR 14-17).

### 4. Discussion

In this double-center observational study, epidemiology and clinical characteristics of 291 COVID-19 patients in Hunan province were collected and analyzed. Hunan is adjacent to Hubei province, and its provincial capital Changsha is 294 kilometers away from Wuhan, while another city Loudi is about 392 kilometers away from Wuhan. Given the migrant data in previous years and the lockdown of Wuhan on Janurary 23, 2020, Changsha is a city with a high estimated number of imported cases.^8^ Our study attempted to provide some experience of diagnosis and treatment for the health workers in other areas. To further show the characteristics in patients with different disease severity, we categorized the 291 patients into mild group (10.0%), moderate group (72.8%) and severe/critical group (17.2%) for statistical analysis.

Current studies have confirmed SARS-CoV-2 as a new branch of the β-coronavirus.^9^ Similar to other β-coronavirus like Severe Acute Respiratory Syndrome Coronavirus and Middle East Respiratory Syndrome Coronavirus, SARS-CoV-2 has shown person-to-person transmission ability since its outbreak in December 8, 2019. The basic reproductive number (R_0_ 3.77) of the COVID-19 estimated by several epidemic prediction models seems to be higher than SARS.^10-12^ After the fast increase in the number of patients in Wuhan during the middle of January, COVID-19 spread to the other provinces rapidly such as neighbor province Hunan because of the nearby location, developed traffic and large migrant population.^11^ In our study, none of 291 patients had a history of direct exposure to Huanan seafood market, but most patients (86.6%) had an indirect exposure history within 14 days before symptoms onset, including being to Wuhan (41.6%), having contact with people came from Wuhan (14.1%) and having contact with local diagnosed patients (38.5%). Our study also showed the proportion of patients who had been to Wuhan in severe/critical group (48.0%) and moderate group (43.4%) were higher than mild group (17.2%). Besides, both the two death cases in our study had a travel history in Wuhan while they both used to be in good health. These showed the infectivity and transmission intensity of the virus, especially in the first or second generation of transmission, and may indicate that the virulence of the virus will decrease after limited generations of transmission.^10,12^ Therefore doctors in areas outside Wuhan should be more cautious in clinical decision making when the patient have a recent history of exposure in Wuhan, including Wuhan residents and those who recently traveled to Wuhan before disease onset. The proportion of family clusters infection in our study was 39.2% which was lower than other studies.^3^ Besides, none of our patient was medical staff while 1080 medical staff were infected in Wuhan until February 11, 2020.^2^

For demographic and clinical characteristics, 53.6% patients aged between 15-49 years, followed by the 50-64 years age group (27.5%). In severe/critical group, 32.0% and 40.0% patients aged from 50 to 64 and over 64 respectively, consistent with report of Guan et al.^13^ Female and male patients both accounted for half in all three groups. The median time from disease onset to first admission in our study was 5.0 days. Similar to the recent publications,^13-16^ our data also showed that fever, cough and fatigue were the most common symptoms. The severe/critical group patients were more likely to have two or more symptoms at admission. While most symptoms were more frequently seen in severe/critical group, nausea or vomiting were more common in mild group, which may indicate different body responses to SARS-CoV-2 infection in patients with different health state and immune system defensive characteristics. 58.0% patients in severe/critical group had at least one underlying disease. In concert with the study of Guan et al,^13^ Bases on these findings, we suggest clinicians pay more attention to and closely observe patients with multiple symptoms and underlying diseases to prevent disease deterioration.

For laboratory inspection, blood routine test showed 36.1% and 22.7% of patients had leukopenia and lymphopenia respectively. Interestingly, eosinopenia was detected in more than 50% patients, especially in the severe/critical group (74%), which was not emphasized in previous studies.^17^ This indicate that eosinophils could assist in the diagnosis and severity assessment of COVID-19. Inconsistent with the other research^10^, elevated D-dimer was observed in majority of patients (83.5%) in our study, and its level was significantly higher in severe/critical group. Consistent with the research of Guan,^13^ Myocardial damage and elevated liver enzyme were not common and mainly happened in severe/critical group. Besides, Obvious disorder of renal function and electrolyte were relatively rare in our patients. 77.6% patients with available coagulation function test result had elevated fibrinogen on admission, and the proportion in severe/critical group was even higher, which have not been observed in other studies. In the early stages of SARS patients, researchers also found an increase in fibrinogen.^18^ Previous studies had shown that SARS-CoV 3a protein can up regulate the expression of fibrinogen in lung epithelial cells.^19^ We speculate in the lung inflammation caused by SARS-CoV-2 or the secondary systemic inflammation, the activated body stress system may lead to the increase of fibrinogen. Another interesting phenomenon was that considerable proportion of patients had positive results in Mycoplasma pneumonia or Chlamydia pneumonia antibody tests, which indicate Mycoplasma pneumonia or Chlamydia pneumonia co-infection, which was higher than other research. Previous report indicated that Mycoplasma fermentans enhanced the cytotoxicity against Vero E6 cells infected with SARS-CoV.^20^ This indicate COVID-19 patients co-infected with the two types of pathogens may lead to more severe state, thus clinicians need pay attention to the screening of these two pathogens in these patients.

Compared with the patient confirmed as COVID-19 in Wuhan^14-16^, our study showed that patients in Hunan had relatively higher discharge rate and lower mortality. In our study, 82.8% patients were prone to a mild or moderate type and 54.6% patients were discharged at the end of follow-up. The length of stay in hospital ranged from 5 to 25 days in discharged patients. 2 of 291 (0.7%) patients died during hospitalization. One death case was a 64 years old man in moderate group without any underlying disease. He had a fever for 3 days before admission and was treated by antiviral therapy including lopinavir and ritonavir tablets. The other death case was a 58 years old man in severe/critical group also without any underlying disease, but had symptoms of fever, cough, dyspnea and fatigue for 7 days before admission. Laboratory tests detected a decreased level of WBC (2.26*10^9^/L), lymphocyte (0.40*10^9^/L) and eosinophils (0.00*10^9^/L), and elevated level of ALT (48.5U/L) and AST (82.1U/L) in this patient. Pulmonary CT of both patients showed progressive patchy consolidation of bilateral lobes. Although there has been a lack of evidence-based specific antiviral drugs, almost all patients in this study received antiviral therapy (96.6%), lopinavir and ritonavir tablets (75.9%), recombinant human interferon α2b (45.4%) were the most commonly used treatment. Besides, chloroquine phosphate was reported to have apparent efficacy and acceptable safety against COVID-19 in a multicenter clinical trials^21^ and had just been included in the latest edition of the guidelines for China. According to this guideline, two patients in the study were given chloroquine for antiviral therapy. However, the safety and efficacy of antiviral therapies used in COVID-19 patients need further studied.

Our study provided more information about epidemiology and clinical profiles of COVID-19 in adjacent area around Hubei Province. It is hoped that our study may provide the basis for the epidemiology related measures of patients in covid-19 import area as well as for clinicians to make medical decisions.

There are several limitations in our study. Firstly, due to the limitations of the retrospective study, laboratory examinations were performed according tothe clinical care needs of the patient, thus some patients’ laboratory exam results were incomlpeted. Secondly, given the short observation period, nearly half of our patients were still receiving treatment in hospital at the end of our follow-up and we could not decide the mortality and prognosis of the whole case series. Moreover, it is difficult to distinguish the specific efficacy of one single drug as various treatment were applied simultaneously, and the guideline about diagnosis and treatment of COVID-19 were updated frequently. Therefore, the treatment experience in our study should be carefully thought when treating patients in different places and circumstances, and further researches are needed to verify the the safety and efficacy.

## 5. Conclusion

In this double-center observational study of 291 hospitalized patients with confirmed COVID-19 in Hunan, a province adjacent to Hubei, 86.6% patients had indirect exposure history. The proportion of patients who had been to Wuhan in severe/critical group and moderate group were higher than mild group. Clinical characteristics of patients in this study were different from patients in Wuhan.

## Data Availability

The datasets used and/or analyzed during the current study are available from the corresponding author on reasonable request.

## Ethics approval and consent to participate

Data collection and analysis of cases and close contacts were determined by the National Health Commission of the People’s Republic of China (PRC) to be part of a continuing public health outbreak investigation and were thus considered exempt from institutional review board approval. Oral consent was obtained from all patients.

## Acknowledgements

We thank all the patients for participating in the present study.

## Author Contributions

Conceived and designed the experiments: Hong Luo and Zhiguo Zhou. Collected Clinical data: Zhilan Yin, Fang Zheng, Yanhua Qing, Dixuan Jiang, Jiyang Liu, Yuanlin Xie, Qi Zuo, Honghui Li, Hong Peng, Yan Chen, Jun He, Jianlei Lv, Analyzed the data: Shuizi Ding, Cheng Lei, Xu Chen. Wrote the text of the main manuscript: Danhui Yang, Shuizi Ding, Xu Chen, Zhiguo Zhou, Hong Luo, Ping Chen. Prepared the tables and figures: Cheng Lei, Shuizi Ding, Xianglin Zhou, Zhilan Yin. All authors reviewed and revised the manuscript.

## REFERENCES

1. NHCPRC. National Health Commission of thePeople’s Republic of China home page: Coronavirus disease 2019 (COVID-19). 2020. http://www.nhc.gov.cn.

2. Novel Coronavirus Pneumonia Emergency Response Epidemiology T. The epidemiological characteristics of an outbreak of 2019 novel coronavirus diseases (COVID-19) in China. Zhonghua Liu Xing Bing Xue Za Zhi 2020; 41(2): 145–51.

3. Xu XW, Wu XX, Jiang XG, et al. Clinical findings in a group of patients infected with the 2019 novel coronavirus (SARS-Cov-2) outside of Wuhan, China: retrospective case series. Bmj 2020; 368: m606.

4. Tian S, Hu N, Lou J, et al. Characteristics of COVID-19 infection in Beijing. J Infect 2020.

5. Lin L, Li TS. Interpretation of Guidelines for the Diagnosis and Treatment of Novel Coronavirus (2019-nCoV) Infection by the National Health Commission (Trial Version 5). Zhonghua Yi Xue Za Zhi 2020; 100(0): E001.

6. Zhu N, Zhang D, Wang W, et al. A Novel Coronavirus from Patients with Pneumonia in China, 2019. The New England journal of medicine 2020; 382(8): 727–33.

7. Ooi GC, Khong PL, Mller NL, et al. Severe acute respiratory syndrome: temporal lung changes at thin-section CT in 30 patients. Radiology 2004; 230(3): 836–44.

8. Wu JT, Leung K, Leung GM. Nowcasting and forecasting the potential domestic and international spread of the 2019-nCoV outbreak originating in Wuhan, China: a modelling study. Lancet 2020; 395(10225): 689–97.

9. Lu R, Zhao X, Li J, et al. Genomic characterisation and epidemiology of 2019 novel coronavirus: implications for virus origins and receptor binding. Lancet 2020; 395(10224): 565–74.

10. Li Q, Guan X, Wu P, et al. Early Transmission Dynamics in Wuhan, China, of Novel Coronavirus-Infected Pneumonia. The New England journal of medicine 2020.

11. Yang Y, Lu Q, Liu M, et al. Epidemiological and clinical features of the 2019 novel coronavirus outbreak in China. 2020: 2020.02.10.20021675.

12. Read JM, Bridgen JR, Cummings DA, Ho A, Jewell CP. Novel coronavirus 2019-nCoV: early estimation of epidemiological parameters and epidemic predictions. 2020: 2020.01.23.20018549.

13. Guan WJ, Ni ZY, Hu Y, et al. Clinical Characteristics of Coronavirus Disease 2019 in China. The New England journal of medicine 2020.

14. Huang C, Wang Y, Li X, et al. Clinical features of patients infected with 2019 novel coronavirus in Wuhan, China. Lancet 2020; 395(10223): 497–506.

15. Wang D, Hu B, Hu C, et al. Clinical Characteristics of 138 Hospitalized Patients With 2019 Novel Coronavirus-Infected Pneumonia in Wuhan, China. Jama 2020.

16. Chen N, Zhou M, Dong X, et al. Epidemiological and clinical characteristics of 99 cases of 2019 novel coronavirus pneumonia in Wuhan, China: a descriptive study. Lancet 2020; 395(10223): 507–13.

17. Zhang JJ, Dong X, Cao YY, et al. Clinical characteristics of 140 patients infected with SARS-CoV-2 in Wuhan, China. Allergy 2020.

18. Sun W, Li ZR, Shi ZC, Zhang NF, Zhang YC. Changes in coagulation and fibrinolysis of post-SARS osteonecrosis in a Chinese population. Int Orthop 2006; 30(3): 143–6.

19. Tan YJ, Tham PY, Chan DZ, et al. The severe acute respiratory syndrome coronavirus 3a protein up-regulates expression of fibrinogen in lung epithelial cells. J Virol 2005; 79(15): 10083–7.

20. Mizutani T, Fukushi S, Kenri T, et al. Enhancement of cytotoxicity against Vero E6 cells persistently infected with SARS-CoV by Mycoplasma fermentans. Arch Virol 2007; 152(5): 1019–25.

21. Gao J, Tian Z, Yang X. Breakthrough: Chloroquine phosphate has shown apparent efficacy in treatment of COVID-19 associated pneumonia in clinical studies. Biosci Trends 2020.

